# Risk assessment of vector-borne disease transmission using spatiotemporal network model and climate data with an application of dengue in Bangladesh

**DOI:** 10.1101/2020.09.16.20195578

**Authors:** Mahbubul H Riad, Lee W Cohnstaedt, Caterina M Scoglio

**Affiliations:** Electrical and Computer Engineering Department, Kansas State University, Manhattan, KS, USA; Arthropod-borne Animal Diseases Research, United States Department of Agriculture, Manhattan, KS, USA

## Abstract

Vector-borne disease risk assessment is crucial to optimize surveillance, preventative measures (vector control), and resource allocation (medical supplies). High arthropod abundance and host interaction strongly correlate to vector-borne pathogen transmission. Increasing host density and movement increases the possibility of local and long-distance pathogen transmission. Therefore, we developed a risk assessment framework using climate (average temperature and rainfall) and host demographic (host density and movement) data, particularly suitable for regions with unreported or under-reported incidence data. This framework consisted of a spatiotemporal network-based approach coupled with a compartmental disease model and a non-homogeneous Gillespie algorithm. One-month and two-month lagged temperature and rainfall data have been used to develop the correlation of climate data with vector abundance and host-vector interactions. This correlation can be expressed as vectorial capacity— a parameter, which governs the spreading of infection from an infected host to a susceptible via vectors. As an example, the novel risk assessment framework is applied for dengue in Bangladesh. Vectorial capacity is inferred for each week throughout a year using average monthly temperature and rainfall data, while the whole country is divided into some spatial locations (upazilas). Long-distance pathogen transmission is expressed with human movement data in the spatiotemporal network. We have identified the spatiotemporal suitability of dengue spreading in Bangladesh as well as the significant-incidence window and peak incidence period. Analysis of yearly dengue data variation suggests the possibility of a significant outbreak with a new serotype introduction. The outcome of the framework comprises of weather-dependent spatiotemporal suitability maps and probabilistic risk maps for spatial infection spreading. This framework is capable of vector-borne disease risk assessment without historical incidence data and can be a useful tool for preparedness with accurate human movement data.

## Introduction

In modern times, threats posed by infectious diseases have become a significant public health concern due to the increasing connectivity [1]. Emerging infectious diseases (e.g., H7N9, H5N1, and Ebola) as well as endemic diseases (e.g., dengue, chikungunya, measles), pose a severe threat to human health and life [2]. Some infectious diseases have high mortality and morbidity rates (Ebola), and some of these diseases lack treatments or vaccines (dengue) [3, 4]. Infectious diseases are creating pandemics due to globalization. For example, 2019 had experienced major dengue outbreaks in many countries in the world, including southeast Asia and Latin America. Other emerging and endemic infectious diseases had also shown to spread rapidly across the globe in recent times. Therefore, accurate risk assessment of disease outbreak is very important for preparedness in this modern world. Risk has been defined as disease development probability within an individual in a specified time interval by medical epidemiologists and health organizations [5]. Risk is also defined as the potential adverse consequences of unwanted phenomena (disease/event)to human life, health, property, or the environment [6]. Accurate risk assessment models have the potential to improve epidemic prevention and control capabilities.

Due to the complexity in vector-borne diseases spreading, often risk is associated with vector or host suitability, the basic reproduction number, vectorial capacity, vector prevalence, or incidence history [5, 7–9]. One method to assess the risk is by detecting disease outbreaks from surveillance data— a retrospective approach, which has a limitation of allowing enough time for preparedness [10]. However, some areas have limited resources for the surveillance system. Especially in developing countries, collected data may not be adequate as most people chose not to use medical facilities unless they have severe conditions. Therefore, disease surveillance data-dependent risk assessment is not always efficient with unreported or under-reported incidences.

Therefore, researchers have developed other risk assessment methods to overcome the problem with retrospective methods. The impact of climate change on the vector survival, suitability and pathogen transmission has been assessed for vector-borne diseases in numerous research [11–19]. Unfortunately, very limited researches have included spatial and temporal heterogeneity of the weather conditions, population demography, and movement information in the risk assessment models.

In this context, we develop a risk assessment framework incorporating the aforementioned significant elements for vector-borne diseases. In this paper, we focus on the mosquito-borne diseases to demonstrate the risk assessment framework as they comprise the majority of vector-borne diseases. This paper has three major contributions. The first contribution is the formulation of a spatiotemporal network-based risk assessment framework by incorporating climate data and demographic information, especially for regions with unreported or under-reported incidence data. The second contribution is deriving a spatiotemporal suitability map of competent mosquito species in the disease transmission with only temperature data. The third contribution is the development of spatial risk maps for disease transmission, showing the relative risk of each location compared to others. Additionally, the identification of the significant-incidence window and peak incidence period is performed by comparing simulation results with data (when available). A serotype analysis is conducted to identify contributing factors for the year-to-year difference in incidence data. This novel risk assessment method is capable of incorporating both human movement and contact patterns as well as impacts of weather factors in human-mosquito interaction.

Finally, an application of the novel framework is presented for dengue spreading in Bangladesh. A spatiotemporal network is developed for human movement in Bangladesh using demographic information, one-month, and two-month lagged climate (temperature and rainfall) data. A map for the spatiotemporal suitability of human-mosquito interaction as well as spatial dengue transmission risk maps are obtained from simulation results. Simulation results matches closely with the significant-incidence window and peak incidence period with Bangladesh dengue transmission dynamics. The year-to-year data variability shows a correlation with the dominant serotype. The combined knowledge obtained from the framework (i.e., significant-incidence window, the peak incidence period, risk map, and spatiotemporal suitability map) provides a guideline to public health personnel in prioritizing spatiotemporal resource allocation to reduce/prevent dengue transmission. Risk maps are developed incorporating generalized human movement data in the spatiotemporal network and have the adaptability to include actual and accurate movement data.

## Materials and method

### Risk assessment framework

Our novel risk assessment framework couples a spatiotemporal network-based approach with a compartmental disease model and a spatiotemporal spreading algorithm. The risk assessment framework has five different components. They are as follows-

- Compartmental model
- Pathogen transmission model with climate data
- Spatiotemporal network
- Spatiotemporal spreading algorithm
- Risk calculation

Each of these components is described in subsequent parts of this section.

#### Compartmental model

Compartmental models express transitions of the host population from one disease state/compartment to another [20]. These compartments are, for example, susceptible, exposed, infectious, recovered, removed, vaccinated, and alert. Some parameters govern inter-compartmental transitions. The transition rate from susceptible to exposed/infected compartment (transmission rate) is the most crucial parameter for the vector-borne disease model. The transmission rate is correlated with climate/weather dependent factors such as vector abundance and host-vector interactions. Therefore, we incorporate climate data in the transmission rate for the developed risk assessment framework.

#### Pathogen transmission model with climate data

The transmission rate for vector-borne diseases has a complicated relationship with the environment and the host. For example, mosquito abundance and their interaction with the host population cause the transition from susceptible to infected (or exposed) states. When the mosquito population is the vector for disease, temperature, and rainfall data are used to develop the correlation between mosquito abundance and their interaction with the host population. This relation can be expressed as *vectorial capacity* — a parameter governing the spread of infection from an infected to a susceptible host via vectors. Vectorial capacity is given as

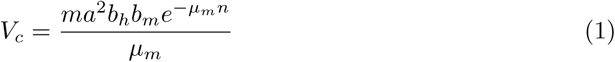

where the vector parameters used are 1) the average daily vector biting rate (*a*), 2) the probability of vector to human transmission per bite (*b*_*h*_), 3) the probability of human to vector infection per bite (*b*_*m*_), 4) the duration of the extrinsic incubation period (*n*), 5) the vector mortality rate (*µ*_*m*_), and 6) mosquito vector density with respect to the host (*m*) [21–23]. These parameters are specific for the mosquito species and the concerned disease. We choose *Aedes aegypti*, one of the most competent mosquito species in transmitting dengue, Zika, chikungunya, yellow fever, and other severe diseases to model the transmission rate/vectorial capacity. Except for mosquito vector density with respect to the host, all other parameters in equation 1 can be calculated empirically using spatiotemporal temperature data [23]. The following empirical formulas are used to calculate temperature-dependent parameters, where *T* is the temperature in degree Celsius.

1. **Biting rate (a):** Liu-Helmersson et al. and Scott et al. developed the following empirical equation from numerous experimental data to model the relationship between temperature and average blood meal frequency of female *A*. *aegypti* [23, 24].

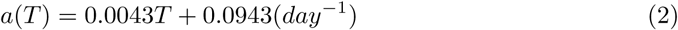
2. **Probability of vector to human transmission per bite (***b*_*h*_): The empirical equation for the probability of human infection was expressed with the following thermodynamic function [23, 25].

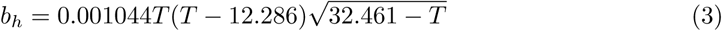

for (12.286^°^*C < T <* 32.461^°^*C*)
3. **Probability of human to vector infection per bite (***b*_*m*_): Lambrechts et al. derived the relationships between temperature and the probability of infection based on empirical data for several *A*. *aegypti* -borne diseases. [26].

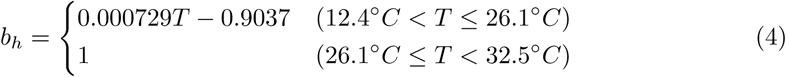
4. **Duration of the extrinsic incubation period (n):** An exponential function was used to fit experimental data for the extrinsic incubation period [26, 27].

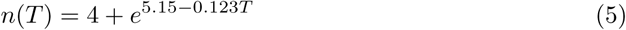
5. **Vector mortality rate (***µ*_*m*_): Yang et al. developed a 4^*th*^ order polynomial equation fitting experimental data to model the mortality rate with temperature [28].

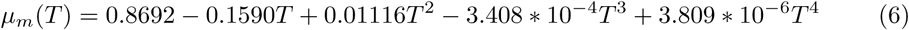
6. **Mosquito vector density with respect to the host (*m*):** The mosquito vector density with respect to the host mostly depends on the rainfall. Therefore, in this article, this parameter is expressed as proportional to the weekly average rainfall [29]. We have normalized the average weekly rainfall in each location before using it in the vectorial capacity model.

#### Spatiotemporal network

To account for the spatiotemporal heterogeneity of the disease transmission risk with changing weather conditions, we propose a spatiotemporal network. The spatiotemporal network is developed using host demographic information such as population density, distribution, and movement. In this network, nodes represent individuals within spatially homogeneous locations, and links represent movements within and between these locations. The network is spatially explicit and has multiple temporal realizations to represent heterogeneities in weather conditions with time and space. This network is then combined with a spatiotemporal spreading algorithm to simulate the spatiotemporal transmission of the infection. The network is periodically updated to reflect the changing weather conditions in the spatiotemporal spreading algorithm.

#### Spatiotemporal spreading algorithm

Nodes influence each other through statistically independent pairwise interactions in most network-based models. Sahneh et al. developed the *generalized epidemic modeling framework* (GEMF) for stochastic spreading processes over complex networks based on these independent pairwise interactions [30, 31]. GEMFsim tool was later developed for numerical simulation of GEMF-based models by implementing the Gillespie algorithm [30]. The combined state of all nodes in a network can be described as a random variable *X*_*N*_ (*t*) = [*x*_1_(*t*), *x*_2_(*t*), …, *x*_*i*_(*t*)], where *x*_*i*_(*t*) is the state (compartment) of node *i* at time *t*. The transition time from one state to another is expressed as an exponential distribution with a transition rate *σ*_*n*_(*x*_*n*_*→ J*), where *J* is the destination state after the transition. This transitions can be node-based (dependent only on the node state *x*_*i*_(*t*)) or edge-based (dependent on the combined network state *X*_*N*_ (*t*)).

After a transition occurs, the combined network state will change, and therefore edge-based transition rates will change. However, node-based transition rates remain constant. GEMFsim accounts for changes in transition rates due to the change in the combined network state. However, GEMFsim does not account for the temporal variation of the transition rates due to external factors (weather conditions or human activities).

The temporal variability of the transition rate is very crucial for simulating vector-borne disease transmission. Therefore, it is required to adapt the Gillespie algorithm in GEMFsim to account for the changing rates. In this article, we incorporate the non-homogeneous Gillespie algorithm in the GEMFsim, which works for exponential event distributions and non-constant transition rates [4, 32, 33]. The modified spreading algorithm is capable of periodically changing transition rates to reflect the temporal heterogeneity of vector-borne disease transmission.

#### Risk calculation

As the spreading process in our spatiotemporal network is highly stochastic, we need to perform an adequate number of simulations. We keep track of each node’s status and count the numbers of simulations in which a particular node is infected. This count is later used to calculate the risk of the spatial disease spreading. The formula for risk calculation is

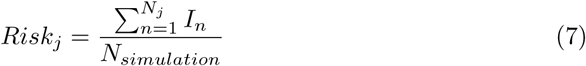

where *Risk*_*j*_ is the spreading risk in location *j, I*_*n*_ is the number of simulations where node *n* is infected, *N*_*simulation*_ is the total number of simulations, *N*_*j*_ is the number of nodes (individuals) in *j*^*th*^ location. The calculated risk is normalized for comparison with the risk of different spatial locations.

### Application of the risk assessment framework for Bangladesh dengue incidence

Dengue is transmitted by *Aedes* mosquitoes in tropical and subtropical regions and may cause a wide range of manifestations from asymptomatic infections to deaths [34].

Arbovirus transmission is known to be driven by the interplay of sex, age, and travel of individuals. Additionally, the transmission also depends on the type of host community (urban/rural), mosquito abundance, and the use of mosquito control measures [35, 35–37]. Understanding the relative importance of these factors is required for assessing the risk of dengue accurately. There has been a recent dengue outbreak in Bangladesh with a record number of cases, drawing close attention to assess the transmission risk. Bangladesh has a history of dengue incidence dated back to 1960, and a major outbreak occurred in 2000 [38–40]. Since 2000, the Ministry of Health and Family Welfare of the People’s Republic of Bangladesh started recording clinical cases, which are reported annually. Recent urbanization throughout the tropical world has accelerated dengue spreading as*Aedes aegypti* — primary vector for dengue transmission— lives in densely populated human-made environments [41]. Bangladesh is a densely populated country with rapid urbanization, which provides a conducive environment for mosquito populations. Therefore, a risk assessment tool for dengue transmission has become very important in Bangladesh.

Several studies have used the historical time series data for reported dengue incidence. However, getting accurate data on dengue infection is surprisingly difficult. Only 11–32% infected people are likely to have symptoms with just a few being sick enough to require formal medical care [41, 42]. Misdiagnosis and under-reporting are common for cases requiring medical care as well. Therefore, risk assessments based on clinical case counts are not always useful, and may just reflect differences in access to healthcare, diagnostics, and the ability to report cases [43]. Record keeping also requires significant resources, which are not always available in Bangladesh. Therefore, our developed framework, which does not require incidence data, can be a useful tool for assessing the spatial dengue transmission risk and the spatiotemporal suitability in Bangladesh. The adapted framework for Bangladesh dengue spreading is presented below.

#### Compartmental model for dengue

When dengue virus enter into the bloodstream of a susceptible person via infected mosquito bites, the individual becomes exposed to the disease. After a specific time for viral replication, the exposed individual becomes infectious. The infectious individual finally transitions to the removed state after recovery or death. Therefore, there are four specific phases/states concerning the disease. These states are named as susceptible, exposed, infected, and removed, and the model is called *SEIR*. The inter-compartmental transitions are independent Poisson processes with transitions rates expressed in equations 8-10.

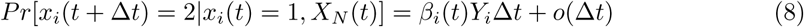

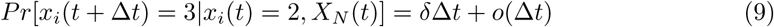

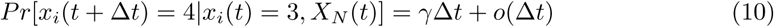

In these equations, *x*_*i*_(*t* + Δ*t*) = 1, 2, 3, and 4 express the probability of node *i* occupying the susceptible, exposed, infected or removed state at time (*t* + Δ*t*), respectively. *X*_*N*_ (*t*) is the combined network state at time *t*. The transition rate from susceptible to exposed state is an edge-based transition, which is also time-variant due to its dependency on weather conditions. We express this time-variant parameter *β*_*i*_(*t*) with vectorial capacity for node *i* at time *t*, which is calculated from spatiotemporal weather conditions. *Y*_*i*_ is the set of infected neighbors of node *i* within the spatiotemporal network at time *t*. The parameter *δ* is the intrinsic incubation rate, which governs the transition from exposed to infected state. The transition from infected to removed state is expressed with the removal rate *γ*. Incubation rate *δ* and removal rate *γ* is node-based transition rates, whose values are assumed equal to 0.17 and 0.14 respectively, and are time-invariant in this work [44, 45]. These values of *δ* and *γ* reflect the means of exponentially distributed parameter values used in the spatiotemporal spreading process.

#### Pathogen transmission model with Bangladesh climate data

Transmission rate *β* is modeled with weekly average temperature and rainfall data in Bangladesh. Climate data are collected from CLIMATE-DATA.ORG for Bangladesh [46]. The upazila level spatial unit is used in this work for network development. Climate data are used to calculate each parameter in the vectorial capacity equation 1. All these parameters, except for mosquito vector density with respect to the host, are calculated from the weekly temperature data. Mosquito vector density parameter is assumed proportional to the weekly average rainfall, and a proportional constant is assumed to reflect a realistic outbreak scenario in Bangladesh.

An urbanization factor is assumed to reflect the suitability of *Aedes* mosquito habitat for dengue transmission. The population density is used to classify the urbanization level of each location. Three urbanization factors are used to reflect backcountry (population density *<* 1000 per square kilometer), rural (1000*≥* population density *<* 3000 per square kilometer), and urban (population density *≥*3000 per square kilometer) locations. The final transmission rate used in simulating dengue transmission is equal to vectorial capacity multiplied by the urbanization factor.

#### Spatiotemporal network

Developing a network is a crucial part of the risk assessment framework. The spatial structure in Bangladesh is as follows-

1. Administrative level 1 — 8 divisions
2. Administrative level 2 — 64 districts
3. Administrative level 3 — 544 upazila

For network development, we have used the administrative level 3 spatial resolution. Therefore, Bangladesh is divided into 544 spatial locations before creating the network. Population data are collected for each spatial location from the City Population website [47]. The population in each location is scaled by 10,000 to reduce the computational burden during simulations.

We assume an Erdos-Renyi network within each upazila, where links are created with a probability of 0.2. Inter-upazila links are created using an exponential dispersion kernel. We use the kernel function *e*^-*kD*^ for link generation, where *k* is a constant, and *D* is the distance between the source and destination location. We choose the value of *k*=0.1 for creating the spatiotemporal network. District-level and division-level human movement, along with the exponential dispersion, are incorporated to reflect the human movement patterns. District-level human movement is incorporated by generating links between the capital city Dhaka and all-district cities. Links are created between Dhaka and all-division cities to include division-level human movements in the network. Fig 1 demonstrates a simplified outline of the network.

**Fig 1.**
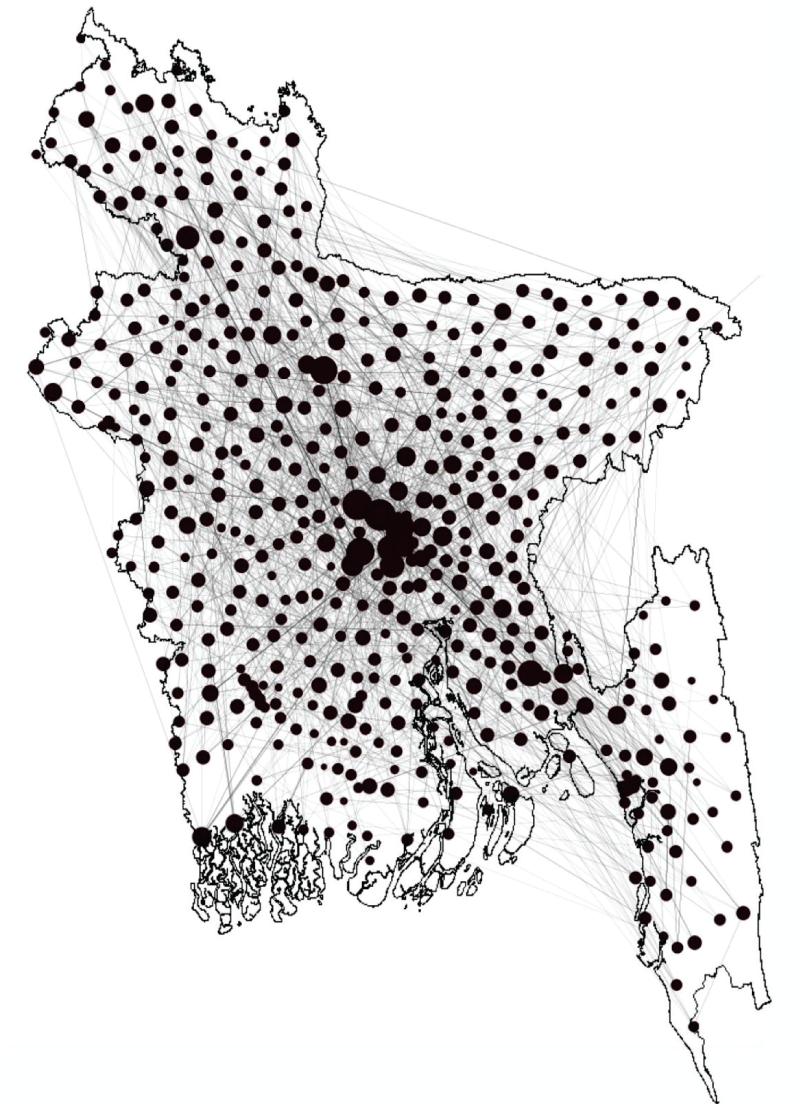
Network for Bangladesh. A simplified diagram for Bangladesh. Each black circle represents the network within an upazila, while lines between circles express the human movement. Circle sizes are scaled according to the human movement (node degree) for that location. Greater circle size indicates a greater amount of human movement flow.

Bangladesh is a small country, having only 147,500 square kilometers of area. However, there is still some spatial and temporal difference between temperature and rainfall throughout the country. This spatiotemporal weather variation is very important for accurately representing dengue transmission. We included a spatiotemporal heterogeneity in the created network due to weather patterns, i.e., pathogen transmission rates on each link. Literature shows the correlation between a one-month and two-month lagged temperature, rainfall, and dengue occurrence in Bangladesh [48]. The heterogeneity in the weather patterns is reflected on a weekly value of transmission rate (*β*) calculated using both one-month and two-month lagged temperature and rainfall data.

We create two instances of the spatiotemporal network for two different outbreak types: first, major outbreaks spreading throughout the whole country, and second, minor outbreaks spreading only within major divisional cities. We explicitly incorporate the district-level human movement for major outbreak scenarios with exponential dispersion kernel. Division level human movement is incorporated for minor outbreak scenarios along with the exponential dispersion. Upon developing the spatiotemporal network, we apply the stochastic spreading algorithm for risk assessment.

## Results and discussion

Assessed risk in this work is a combination of weather-dependent spatiotemporal suitability and risk maps from the network-based model. Therefore, we present our results in the following two sections such as-

- Spatiotemporal suitability of dengue transmission in Bangladesh
- Risk maps for dengue transmission in Bangladesh

### Spatiotemporal suitability of dengue transmission in Bangladesh

Spatiotemporal suitability expresses the spatial and temporal suitability of the vector (mosquito) survival and functioning in pathogen transmission. When comparing dengue epidemic potential over time and space, it is preferable to use the relative vectorial capacity [23]. Relative vectorial capacity is expressed as the vectorial capacity relative to the vector-to-human population ratio and formulated as 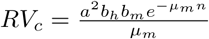. All parameters are temperature-dependent in the relative vectorial capacity definition. A higher relative vectorial capacity indicates a higher potential for the dengue epidemic. We calculate the relative vectorial capacity for *Aedes* mosquito in Bangladesh to infer the suitability of dengue spreading. There exists a threshold value for relative vectorial capacity beyond which *Aedes* mosquitoes can function properly in transmitting dengue infection. A relative vectorial capacity greater than 0.6 indicates the suitability of dengue spreading [23]. The spatiotemporal suitability maps for dengue spreading in Bangladesh are presented in Fig 2 for each month of the year. The relative vectorial capacity is found higher than the threshold value (0.61) for many months of the year in almost all locations (red and yellow regions).

**Fig 2.**
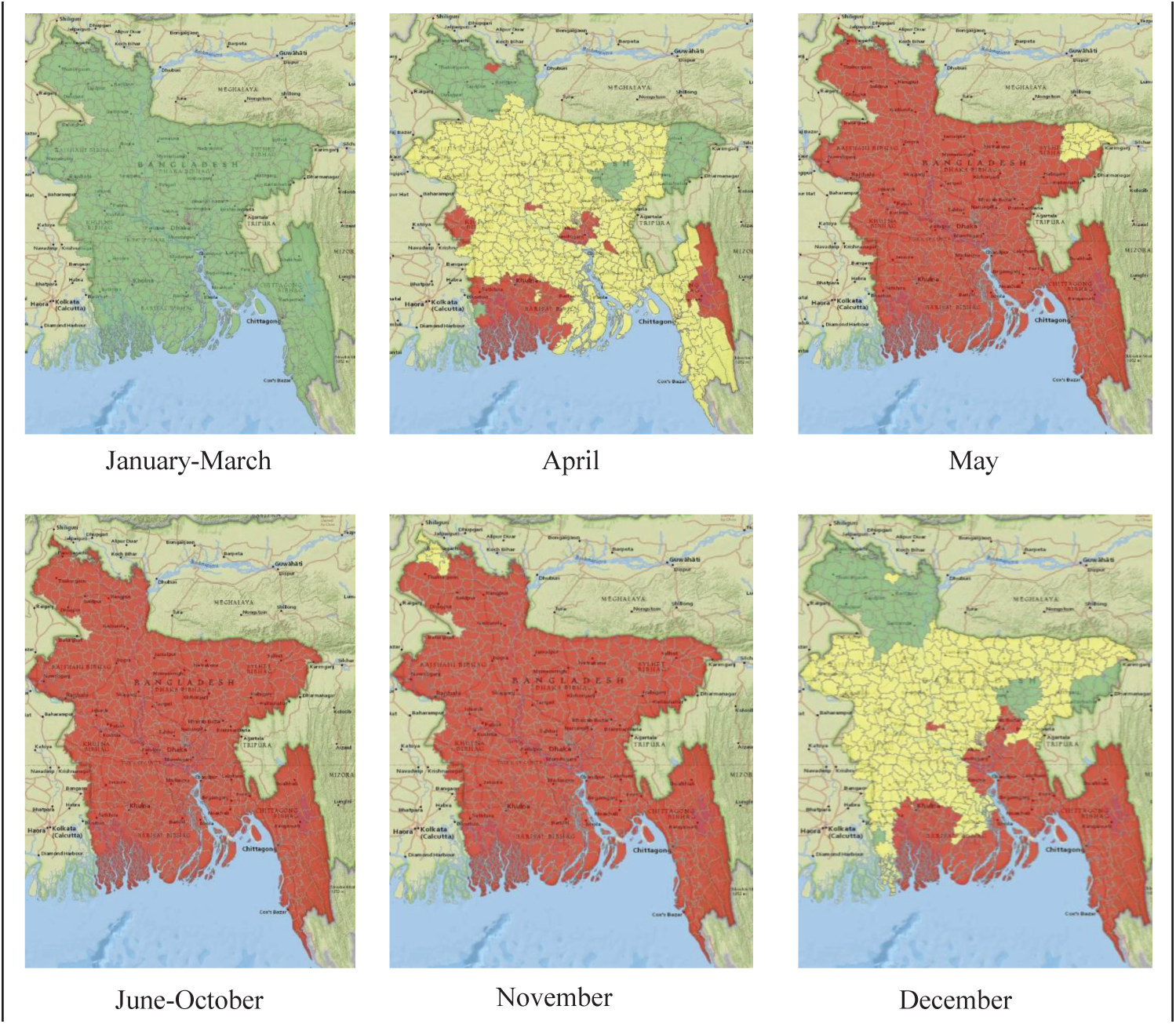
Spatiotemporal suitability maps for dengue transmission in Bangladesh. Spatiotemporal suitability maps for dengue transmission in Bangladesh based on temperature. The map represents suitability in the following manner-green (lowly suitable), yellow (moderately suitable), and red (highly suitable).

It is evident from Fig 2 that months between January and March are poorly suited for dengue transmission. In April, some southern parts of the country, as well as capital city Dhaka, become highly suitable. The other parts of the country, except the north most corner, become moderately suitable. Starting from May, the whole country becomes highly suitable, and the situation remains similar until November. The most northern part of the country becomes lowly suitable while the capital and southern part stays highly suitable in December. After December, the whole country becomes poorly suited again, which continues until April.

Dengue incidence data since 2000 shows cases throughout the year, which supports our results for the suitability of dengue transmission. Bangladesh is a tropical country, which provides a suitable temperature for mosquito survival year-round. However, for temperate regions with widely varying temperatures, mosquitoes may not be able to survive in the coldest months.

### Risk maps for dengue transmission in Bangladesh

The network-based model enables us to simulate the spreading process within the spatiotemporal network described above. Every year, the dengue outbreak in Bangladesh shows a different trend. Some years, cases are reported from most parts of the country, while some years outbreaks are mostly limited within divisional cities. Depending on the spreading, outbreaks are divided into two categories— major outbreaks with widespread dengue cases and minor outbreak with cases mostly in some divisional cities. Therefore, two distinct simulation scenarios are assumed to match the two different outbreak types. Simulations are performed with parameters calculated using both one-month and two-month lagged temperature and rainfall data, which are presented in the subsequent parts of this section.

#### Scenario 1: major outbreak

A major outbreak is defined when dengue cases are widespread throughout the whole country. For major outbreaks, the network is generated with district-level human movements incorporated. Dhaka is the capital of the country; therefore, frequent movements between all district towns are assumed to and from Dhaka. We performed one-thousand iterations for each scenario to account for the stochasticity in the simulation results. Simulations are started with an entirely susceptible population with one infected human in the initial outbreak location. The initial outbreak location is also changed to see the impact of human movements on the transmission risk. Simulation results for one-month lagged climate data, and two initial conditions— Dhaka and Chittagong outbreak starting are presented in Fig 3.

**Fig 3.**
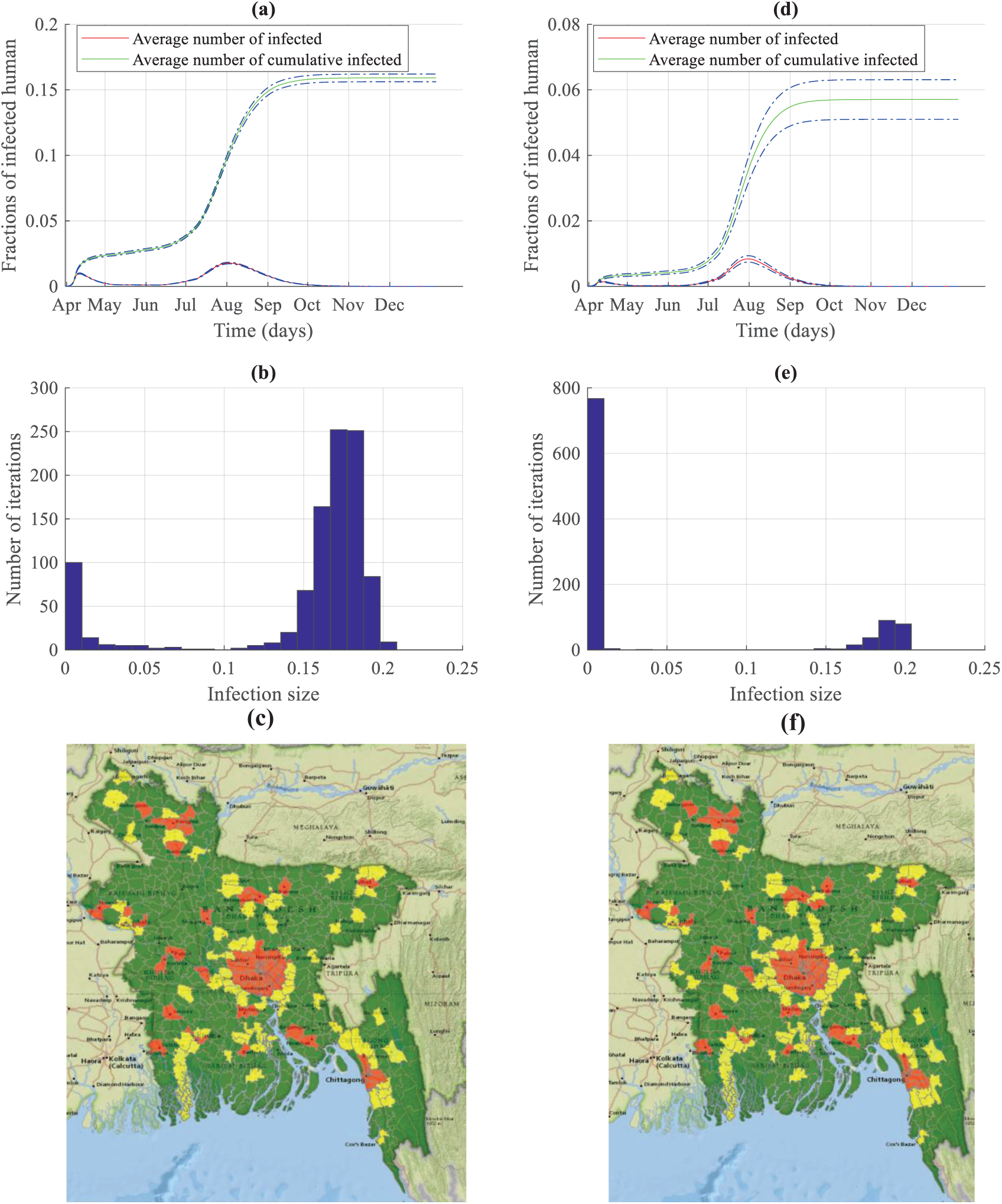
Simulation results and risk maps for dengue transmission in Bangladesh for a major outbreak. The left side panels are results of simulations started in Dhaka, while the right side panels are results of simulations started in Chittagong. Panels (a) and (d) show the dengue transmission dynamics; Panels (b) and (e) present histograms of the number of simulations and infection size; Finally, panels (c) and (f) display risk maps for dengue infection.

Fig 3(a) and 3(d) shows the fractions of infected people and cumulative infected people with 95 % confidence interval when the simulation started in Dhaka and Chittagong, respectively. The curve for the average number of infected shows two peaks in Fig 3(a) and 3(d). The first peak refers to the rapid spreading within the vicinity of the initial outbreak location, and the second peak represents the widespread outbreak. For novel vector-borne diseases, the infections start with a single infected human or a single infected mosquito. If we assume a single infected human started the infection, then the infection will start locally with competent mosquitoes biting the infected person. As mosquitoes have a short flying range, the infection will be local at the beginning, and this is evident from the smaller initial peak. The size of the peak depends on the population size, level of urbanization, and the area of the outbreak location. Being Dhaka densely populated, an outbreak starting in Dhaka will result in a pronounced initial peak due to only the local transmission (Fig 3(a)). We started our simulation in April as it is the first month of high suitability for *Aedes* mosquito within a year for Bangladesh. However, the suitability increases with time, as shown in Fig 2 and the infection starts spreading to distant locations due to human movement as well as higher suitability. Hence, we have another peak in August with numerous cases countrywide. Although Fig 2 shows all locations are highly suitable from May to November, the underlying mosquito dependent transmission rate keeps increasing until August and attains higher values in June-August. Therefore, despite human movements from Dhaka to other locations all year, due to the lower value of transmission rates, no widespread infections start until June. With the higher value of the transmission rate, the number of infected individuals keeps increasing from June and attains its peak in August. Around 16% of the total population in the network becomes infected in this outbreak scenario. Almost 70% of the total infection are observed within the period of July-September. The peak consists of 2.5% of the entire population in our developed network. These infected people during August may require hospital care, which will be a huge burden on the healthcare system. Therefore, proper measures should be taken, and more resources should be allocated for dengue healthcare during July-September. Fig 3(d) shows the dynamics when dengue infection started in Chittagong. Total fractions of cumulative infected people and infected people during peak time are both significantly lower for this outbreak scenario, which can be attributed to the fact that Chittagong is not very densely populated as well as not well connected to the whole country as Dhaka. Therefore, both the initial smaller peak and the second higher peak are smaller compared to the peaks in the Dhaka outbreak scenario. When the infection starts in Chittagong, around 6% of the population becomes infected compared to 16% in the Dhaka outbreak scenario. Therefore, the infection starting location is a crucial determining factor of the extent and the epidemic dynamics.

Fig 3(b) and 3(e) show the histogram with fractions of cumulative infected humans in the Bangladesh dengue network and the number of simulations performed. The axis represents fractions of infected humans in simulation, and the y-axis shows the number of simulations where a particular infection size is obtained. Fig 3(b) shows that almost 80% simulation results in 10-20% infected humans in the representative network when the initial outbreak happens in Dhaka. This accounts for the narrower confidence interval in Fig 3(a). Fig 3(e) express around 20% probability of 10-20% human being infected. This variability in the fractions of infected individuals accounts for the wider confidence interval in the simulation results in Fig 3(d).

Transmission dynamics presented in Fig 3 are obtained using the network with district-level human movement incorporated and one-month lagged temperature and rainfall data. However, when we perform simulations with two-month lagged temperature and rainfall data, the simulation is started in May, and the peaks are a month delayed. The analysis of Bangladesh dengue incidence data since 2000 showed the occurrences of peaks in July-October [38, 39]. Therefore, two-month lagged data also show significant similarity with the actual incidence data. Risk maps are similar for both one-month lagged, and two-month lagged data. Simulation results for two-month lagged data are presented in the S1 Fig.

#### Scenario 2: minor outbreak

Dengue infections are often confined within divisional cities, and a widespread outbreak does not happen. Therefore, we propose another scenario where we explicitly incorporate human movement from Dhaka city to other divisional cities along with the distance-based exponential movement kernel. We call this scenario a minor outbreak as the infection does not spread countrywide. We perform simulations for both one-month and two-month lagged climate data.

The results are presented in Fig 4, when simulations are performed with one-month lagged climate data and the same initial conditions as major outbreak scenarios, namely starting simulations in Dhaka and Chittagong.

**Fig 4.**
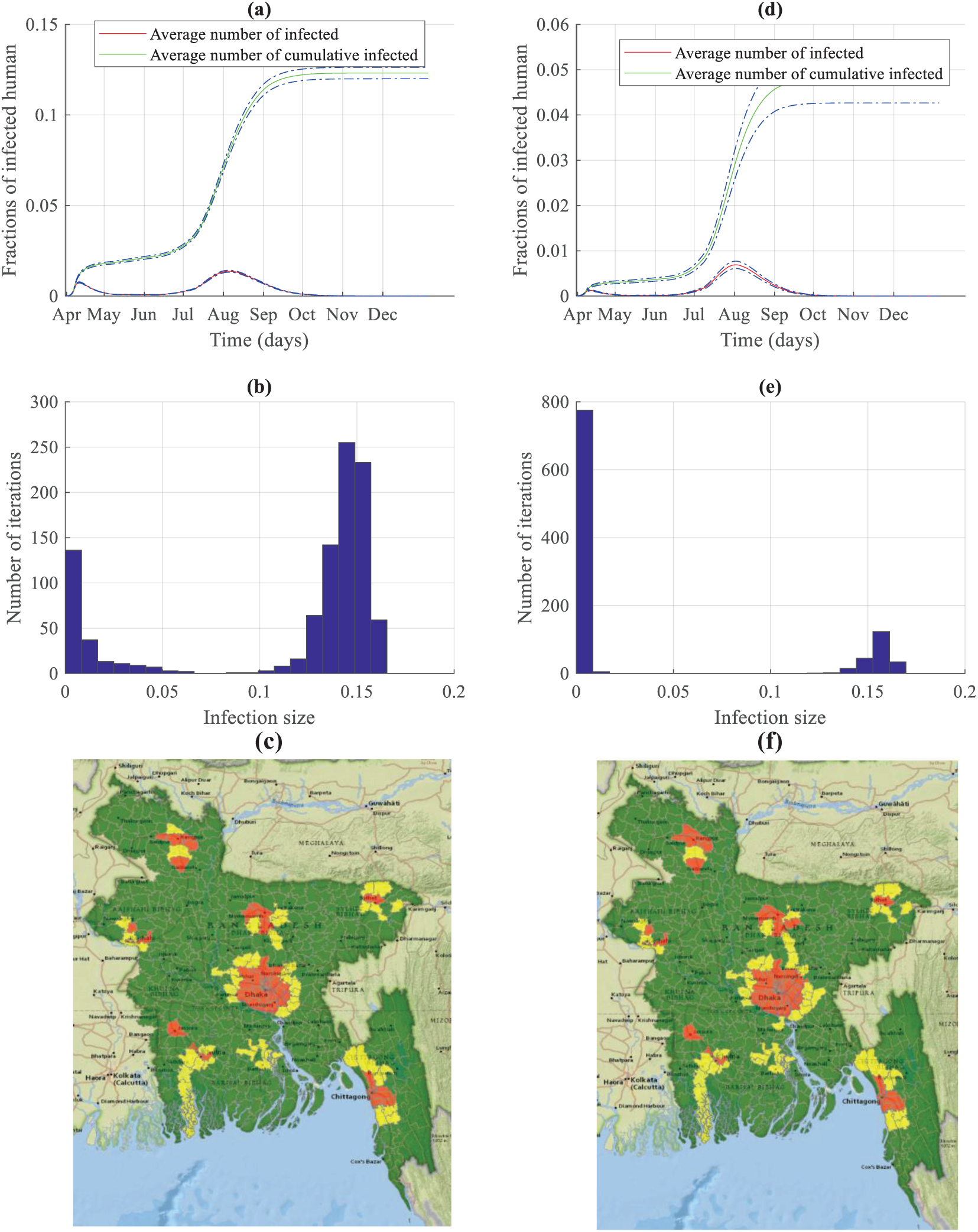
Simulation results and risk maps for dengue transmission in Bangladesh for a minor outbreak. The left side panels are results of simulations started in Dhaka, while the right side panels are results of simulations started in Chittagong. Panels (a) and (d) show the dengue transmission dynamics; Panels (b) and (e) present histograms of the number of simulations and infection size; Finally, panels (c) and (f) display risk maps for dengue infection.

Fig 4(a) and 4(d) show a similar trend as shown in Fig 3(a) and 3(d). For one-month lagged data, a major peak is observed in August with a rapidly increasing infection within July-September. However, comparing Fig 4 and Fig 3, we can see that both the value of the fraction of infected people during the peak infection time and the whole outbreak period are smaller during minor outbreaks.

Human movement can also be considered as a crucial factor in the vector-borne disease spreading, although the pathogen transmission does not happen via direct physical contact. An exposed/infected person may move/travel to a different location and become infectious after reaching the destination. That person can be bitten by a competent local mosquito and may start a local outbreak in the destination location. Therefore, mosquitoes are responsible for the local transmission, while human movement is mostly responsible for long-distance pathogen transmission during a period of higher suitability. The reduction in the number of infections in the minor outbreak scenario can be attributed to the reduced human movement volume within the network than the major outbreak scenario.

It is evident from the comparison between panels (a) and (d) of Figs. 4 and 3 that the major dengue spreading period in Bangladesh is June-September. Finding this time window for significant transmission is very crucial for public health officials. The identification of the significant-incidence window will enable them to take prompt actions during the surge of infection. The major action for controlling a vector-borne disease is always controlling the vectors. Therefore, control measures need to focus mostly on the significant-incidence months to reduce the mosquito population when resources are inadequate for the whole year.

Fig 4(c) and 4(f) show the normalized risk maps for dengue spreading during a minor outbreak. It is evident from the figure that high-risk areas are confined within major division cities and their nearby locations in contrast to Fig 3(c) and 3(f), where many locations throughout the country are at high risk. This reduction in the spreading risk can be attributed to the reduced human movement in the minor outbreak scenario. Therefore, making people aware of the human movement’s impact on long-distance travel through social media or radio/TV broadcasting would help contain the epidemic.

Simulations are also performed for two-month lagged climate data, and all the results are similar to the ones obtained with one-month lagged data. These simulation results are presented in S2 Fig.

Our proposed framework is a generalized risk assessment tool based on climate and demographic data, which can be used for risk assessment, especially in regions with unreported or under-reported incidence data. Risk maps developed in this work are generated with the generalized concept of human movement within Bangladesh. The framework can incorporate more detailed and accurate human movement data. The incorporation of detailed movement data will provide a more accurate assessment of the transmission risk of each location. Once proper and accurate movement data is incorporated in the network, the control measures should be applied to the high-risk areas first, followed by the medium and low-risk areas depending on the availability of resources.

### Serotype analysis

Since 2000, there are dengue cases every year in Bangladesh. However, the number of cases varies from year to year. Importantly, the available data concern only hospitalized and reported cases of dengue. In this section, we show the existence of a correlation between the number of cases and the circulating DENV serotypes. Dengue fever can be caused by any of four genetically related dengue virus (DENV) serotypes (DENV1, DENV2, DENV3, and DENV4) [49]. After recovering from infection with one dengue serotype, a person has immunity against that particular serotype [49]. Unfortunately, the person can be infected again with any of the remaining three dengue serotypes [50]. Subsequent infections often put individuals at a greater risk for severe dengue illnesses than those who have not been previously infected [51]. A bar graph of yearly dengue incidences in Bangladesh with serotypes is presented in Fig 5.

**Fig 5.**
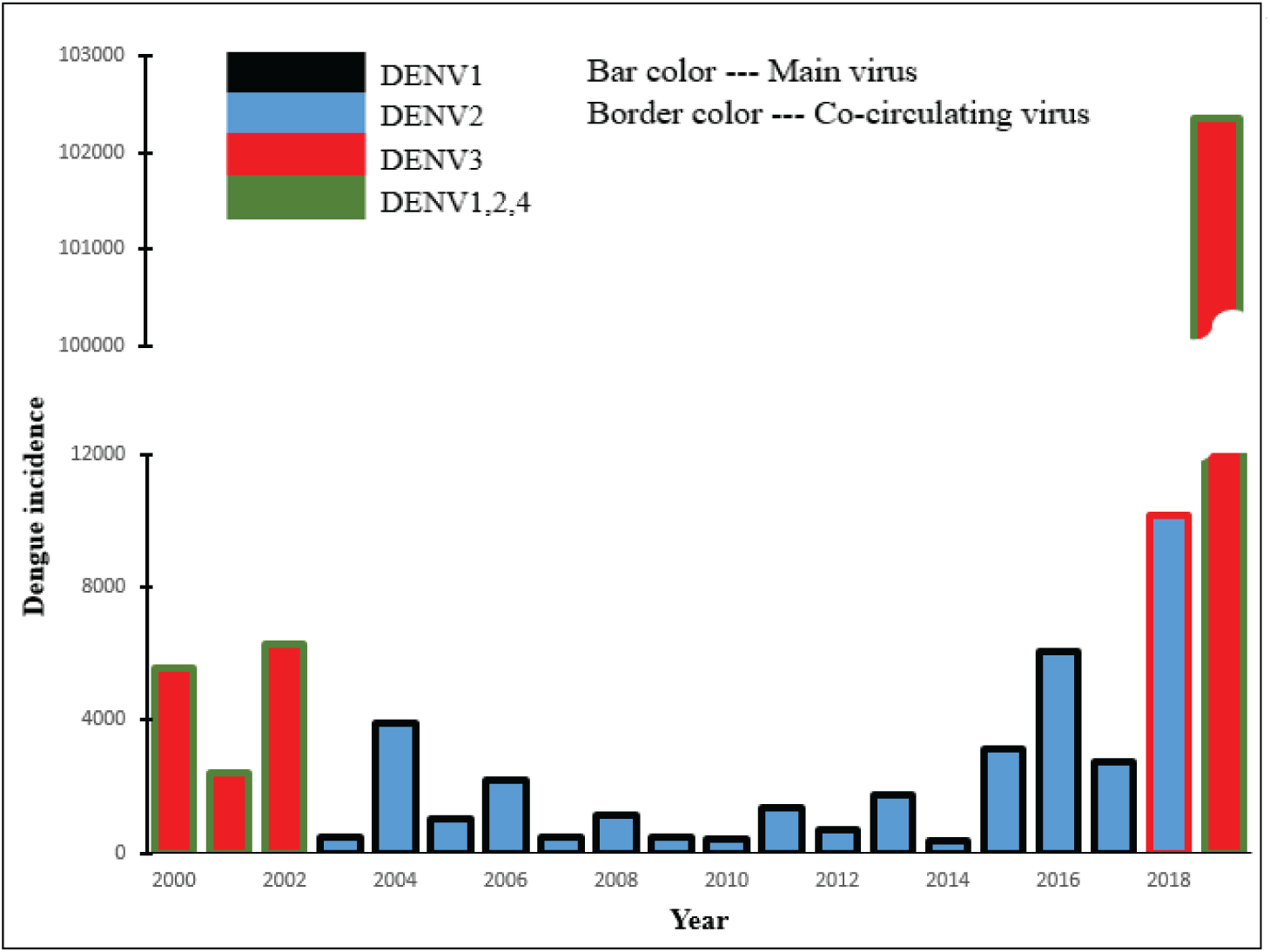
Serotype analysis of dengue spreading in Bangladesh since 2000. The bar chart presents the number of dengue cases with the circulating serotypes each year in Bangladesh. The main bar color represents the dominant serotype, while the border represents other circulating serotypes.

The primary bar color represents the dominant circulating serotype, while the border-color represents the co-circulating serotypes. The x-axis shows the year, and the y-axis shows the number of reported dengue cases in Bangladesh. The bar and border colors are as follows-black for DENV1, blue for DENV2, red for DENV3, and green for DENV1,2,4 together. The height of each bar corresponds to the number of cases in the corresponding year. Fig 5 shows that the dominant circulating serotype was DENV3, with the other three co-circulating, during the years 2000-2002. Within the period 2003-2016, DENV-2 was the dominant serotype, and DENV-1 was co-circulating [50]. The DENV3 reemerging in 2017 resulted in a significant increase in the number of cases during 2018. However, DENV2 was still the dominant circulating serotype in 2018 [50]. If a serotype is circulating long enough within a population, all recovered people may become immune to that specific serotype. An increase in the number of cases in 2018 can be attributed to the reemergence of DENV3. DENV3 became the dominant circulating serotype in 2019, which created an extreme and unprecedented surge in the number of infections. More than a hundred thousand cases were reported in 2019— which is more than double of combined cases in the previous nineteen years— due to a vast susceptible population for DENV3 serotype. Currently, DENV3 is already in circulation in Bangladesh. Neighboring countries of Bangladesh have all four serotypes. Therefore, Bangladesh is always at risk for all four serotypes, including DENV4.

Healthcare personnel should be vigilant to identify dengue patients before the significant-incidence period to identify the circulating serotypes. The introduction of a new serotype will produce a surge in dengue infections with a high probability.

### Peak timing validation

The yearly reported cases varied widely for dengue incidence in Bangladesh. For example, there were 10148 cases in 2018, and more than a hundred thousand cases were reported in 2019 [50, 52]. Therefore, there is high variability in year-to-year dengue cases. From the serotype analysis in the previous section, this 2019 unprecedented increase can be attributed to the DENV-3 circulation. The yearly number of dengue cases is a complex combination of circulating serotypes, the movement patterns of the human population, and the measures taken for mosquito control. For widely varying year-to-year case numbers, we used the peak incidence time and the transmission dynamics to compare our simulation results with the incidence data. Peak incidence happened mostly in August and September in Bangladesh [50]. Simulation results with one-month lagged climate data show incidence dynamics with the major peak in August. Two-month lagged climate data resulted in a peak in September. We compared our simulation with actual incidence data for 2018 and 2019 in Fig 6.

**Fig 6.**
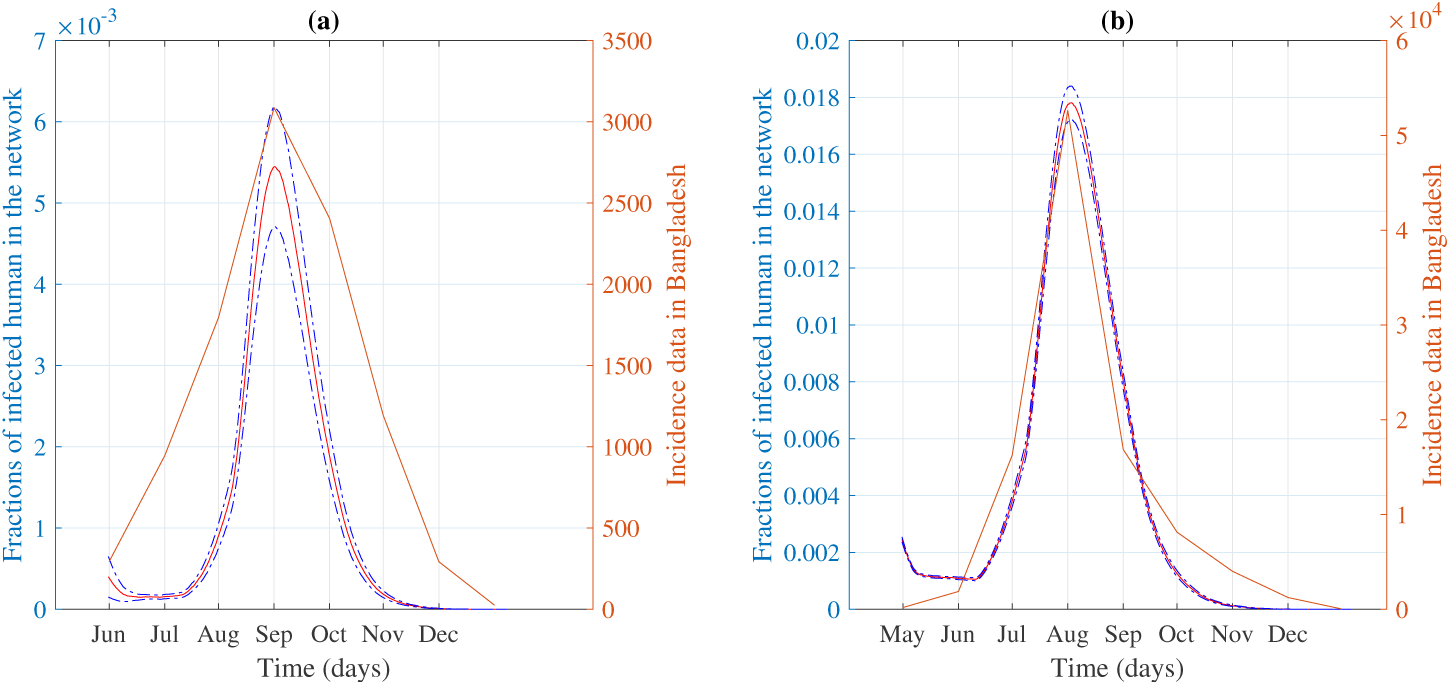
(a) Comparison of peak time from our simulation with incidence data during a minor outbreak in 2018; (b) Comparison of peak time from our simulation with incidence data during a major outbreak in 2019.

In 2018, the peak of the incidence data was observed in September, as shown in Fig 6(a). Simulation results for two-month lagged climate data show a peak in September in accordance with 2018 incidence data in Fig 6(a). The disease dynamics from simulations with one-month lagged climate data shows a similar trend with a coinciding peak for 2019 (Fig 6(b)).

### Application of control measures

The application of control measures decreases the spatial spread as well as the number of dengue cases during an outbreak. Main control measures include spraying insecticide and adopting preventive measures to avoid mosquito bites. To stay away from mosquito bites, one can wear clothes that cover most parts of the body, sleep under bed nets, and use mosquito repellents. Decreasing the probability of mosquito bites per day as well as decreasing mosquito abundance will check the widespread outbreak of dengue. To demonstrate the impact of the control measures, we performed our simulations with multiple reduced transmission rates. Fig 7 shows the effect of a 50% reduction in the transmission rate in the disease dynamics and the corresponding risk map.

**Fig 7.**
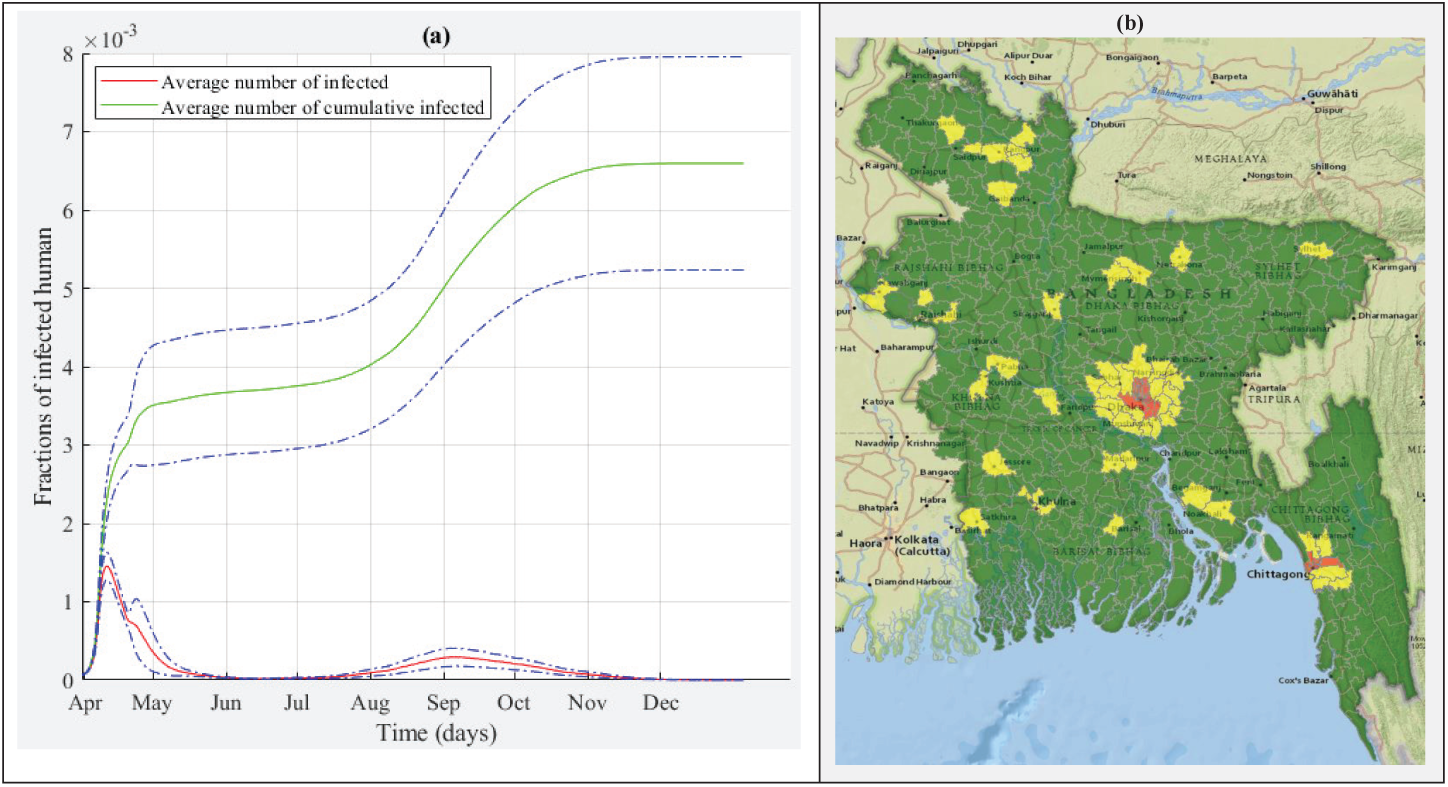
(a) Temporal spreading of dengue with control measures implemented; (b) Spatial risk map of dengue spreading when control measures are applied.

Fig 7(a) shows the disease dynamics after the control measures implemented when the infection started in Chittagong. The fraction of infected individuals is greatly reduced as compared to Fig 4 and 3. Fig 7(b) shows the risk map for dengue transmission after control measures have been applied. The high-risk areas now become confined within the initial outbreak location and some areas in the capital city. The application of control measures reduced both the risk of spatial dengue transmission as well as the total number of cases. Therefore, proper application of control measures, especially during the significant-incidence period identified from our simulation, would be very effective in reducing the epidemic transmission.

## Conclusions

Climate data can be used to develop a generalized risk assessment framework for vector-borne diseases together with demographic data— spatial distribution of individuals and their movement patterns. Therefore, we develop a novel risk assessment framework with a spatiotemporal network model and a non-homogeneous Gillespie algorithm using both climate and demographic data. The assessed risk from this framework is comprised of spatiotemporal suitability maps and spatial risk maps.

Spatiotemporal suitability maps show the spatial and temporal suitability of vector-borne transmission. Spatial risk maps represent the disease transmission risk of each location compared to other locations on the map. This framework also identifies the high risk or elevated risk-months as well as the peak incidence period within a year.

Upon development, the framework is applied to the study of dengue transmission in Bangladesh for major and minor outbreak scenarios. The difference between major and minor outbreaks is defined by different levels of human movement to demonstrate the critical role of human dispersal on widespread pathogen transmission. Reduced human migration throughout the country will reduce the infection spread to divisional cities. We generate the spatiotemporal suitability map and the risk maps for Bangladesh dengue transmission. Simulation results also showed a similar significant-incidence window as well as the peak incidence period with reported dengue incidence data in Bangladesh. Serotype analysis indicates the importance of identifying circulating DENV serotypes before the significant-incidence window. The possibility of a major outbreak is associated with the introduction and reemergence of new DENV serotypes. Simulations of control measure applications, such as mosquito control or other preventive behaviors, have shown a significant decrease in the number of cases as well as risk. The proposed risk assessment tool provides guidelines for public health officials to prioritize resource allocation and control measure application according to the estimated risk.

## Data Availability

Data will be available upon publication

## Acknowledgement

The project was sponsored by the Department of the Army, U.S. Army Contracting Command, Aberdeen Proving Ground, Natick Contracting Division, Ft Detrick, MD (DWFP grant W911QY-19-1-0004), and NSF/NIH/USDA/BBSRC Ecology and Evolution of Infectious Diseases (EEID) Program through USDA-NIFA Award 2015-67013-23818.

## Supporting information

S1 Fig (a) Transmission dynamics of dengue in Bangladesh when the simulation started in Dhaka with two-month lagged climate data and district-level movement network (b) Corresponding histogram (c) Spreading Dynamics of dengue in Bangladesh when the simulation started in Chittagong with two-month lagged climate data and district-level movement network (d) Corresponding histogram

S2 Fig (a) Transmission dynamics of dengue in Bangladesh when the simulation started in Dhaka with two-month lagged climate data and division-level movement network (b) Corresponding histogram (c) Spreading Dynamics of dengue in Bangladesh when the simulation started in Chittagong with two-month lagged climate data and division-level movement network (d) Corresponding histogram

